# Changes in oral risk habits and influencing factors among participants in an oral cancer screening program

**DOI:** 10.1101/2025.02.20.25322636

**Authors:** Pattaranan Munpolsri, Chiu-Wen Su, Hsu-Fei Yang, Tsui-Hsia Hsu, Yen-Yu Chou, Li-Ju Lin, Chao-Chun Wu, Sam Li-Sheng Chen, Amy Ming-Fang Yen

## Abstract

This study examines changes in oral risk habits and identifies factors influencing these changes among participants in a population-based oral cancer screening program to support effective public health interventions. The study included 2,569,920 individuals aged 30 and older who participated in Taiwan’s Oral Cancer Screening Program at least twice between 2010 and 2021. Changes in cigarette smoking and betel quid chewing were assessed between the first and last screenings and categorized as improved, unchanged, or worsened. A logistic regression model evaluated factors associated with habit improvement, including baseline oral habits, sex, age, education, screening adherence, and oral potentially malignant disorder (OPMD) findings.

Among participants, 25.3% improved their oral habits. Baseline habits influenced how OPMD screening results affected behavior change. Among smokers, a positive screening result increased the likelihood of quitting or reducing smoking (adjusted odds ratio [aOR] = 1.18, 95% CI 1.16–1.20). However, among betel quid chewers, whether or not they smoked, a positive screening result was negatively associated with improved habits (aOR 0.79–0.88). Being female, older, college-educated, and regularly attending screenings were positively linked to behavior improvement.

The program led to habit improvements in about one-quarter of participants, particularly older individuals, those with higher education, and frequent attendees. However, a diagnosis of OPMD motivated change only among smokers, not those engaging in both smoking and betel quid chewing, highlighting a lack of awareness in high-risk groups. Strengthening collaboration between health organizations and the screening program could enhance public awareness, improve program effectiveness, reduce oral cancer incidence, and lower long-term healthcare costs.

## 1 Introduction

Oral well-being is a crucial aspect of overall health, and preventive measures are vital in mitigating the risks associated with detrimental oral habits. Various oral risk habits, such as smoking and betel quid chewing, are significant contributors to oral health challenges, including an elevated risk of oral cancers and related conditions. Oral cancer is the most hazardous disease among all types of oral diseases resulting from abnormal cell growth in the oral cavity. Although the specific causes of this abnormality remain unidentified, it is evident that certain habits significantly increase the risk of oral cancers such as smoking, alcohol consumption, betel quid chewing, oral infections, and oral potentially malignant disorders (OPMDs) [1–3]. Oral cancer screening programs can effectively reduce mortality by enabling early detection of oral cancer and OPMDs, allowing for timely referral to appropriate treatment [4–6]. Recognizing the importance of early detection and intervention, oral screening programs are essential tools for promoting awareness of oral health and encouraging habit changes [7]. Participants in these screening programs are expected to modify their habits accordingly to further reduce the risk of developing the disease.

Previous research, whether based on surveys or cohort databases, suggests that factors such as dosage of consumption, educational level, illness, and intention to adjust habits have an impact on the likelihood of success in improving habits. Nevertheless, these investigations are limited to enhancing a singular habit [8–14]. However, in actuality, the majority of individuals with oral risk habits do not possess only one risk habit, and these habits also have an impact on one another [15–21].

This study aims to report the changes in oral risk habits among individuals participating in an oral screening program that targeted those engaged in high-risk oral habits, including cigarette smoking and betel quid chewing. We also investigated factors associated with changes in oral habits, yielding more precise insights into the consequences of harmful modifications. The primary objective of this study is to investigate the changes in oral risk habits and assess the factors linked to the adoption of advantageous oral habits among individuals participating in a population-based oral cancer screening program in Taiwan. This analysis will consider combining two specific oral risk habits, cigarette smoking and betel quid chewing.

## 2 Material and Methods

### 2.1 Study design and setting

This is a retrospective cohort study that uses data from the oral cancer screening program in Taiwan.

### 2.2 Data source and collection

This study utilized data from the Taiwan Oral Cancer Screening Program spanning 2010–2021. Participants aged 30 and older who smoked or chewed betel quid underwent biennial screenings for oral neoplasia to reduce oral cancer incidence and mortality. A screening data monitoring and surveillance center recorded screening, referral, and diagnosis data. Demographic information, oral habits, and screening results were collected at each visit. Chuang (2017) ^[6]^ outlined the coding issues and specifics of this screening program. A structured questionnaire was used to gather demographic data and information on cigarette smoking and betel quid chewing through face-to-face interviews conducted in community and hospital settings. Participants’ oral cavities were visually examined by trained dentists or physicians to identify Oral Potentially Malignant Disorders (OPMD), including oral leukoplakia, erythroleukoplakia, erythroplakia, oral submucous fibrosis, and verrucous hyperplasia. It is recognized that interview-based data collection is prone to biases, including interviewer bias, social desirability bias, and recall bias. To minimize these biases, all interviews were conducted by trained professionals or personnel skilled in interview techniques. For study purposes, data from January 2010 to December 2021 was accessed on November 22, 2022. Information that could identify individual participants during or after data collection was accessible only to Amy Ming-Fang Yen, Ph.D.

### 2.3 Participants and variables

This study uses data of 2.6 million participants from the oral cancer screening program in Taiwan between 2010 and 2021 with at least two times of screening. The data included oral habits change, age (30-45, 46-60, 60+), sex, education level (elementary school, middle and high school, college+), screening times (only two times, more than two times), and OPMDs (positive-, negative finding). The education on oral cancer risks and prevention was done as a part of the screening program to follow up on their effectiveness in which the oral habit changes were collected every time of screening.

### 2.4 Measurement of cigarette smoking and betel quid chewing

This study intends to investigate two habits, cigarette smoking and betel quid chewing. The use of cigarette smoking is categorized into 3 levels: LS: never smoke, cessation, and low degree (<10 years and <20 pieces per day); MS: medium degree (<10 years and ≥20 pieces per day, ≥10 years and < 20 pieces per day); and HS: high degree (≥10 years and ≥20 pieces per day). The betel quid chewing is categorized into 5 levels NB: never chewing, QB: cessation, LB: low degree (<10 years and <20 pieces per day), MB: medium degree (<10 years and ≥20 pieces per day, ≥10 years and < 20 pieces per day), and HB: high degree (≥10 years and ≥20 pieces per day). The combined oral habit changes between the first and the last screening for cigarette smoking and betel quid chewing was rated to improve and not improve (Fig 1a). The improvement of the combination of smoking and betel quid chewing indicates that at least 1 habit being improved with no other habit worsening (Fig 1d). The improvement of smoking and betel quid chewing was defined as shown in Figs 1b and 1c.

**Figure 1.**
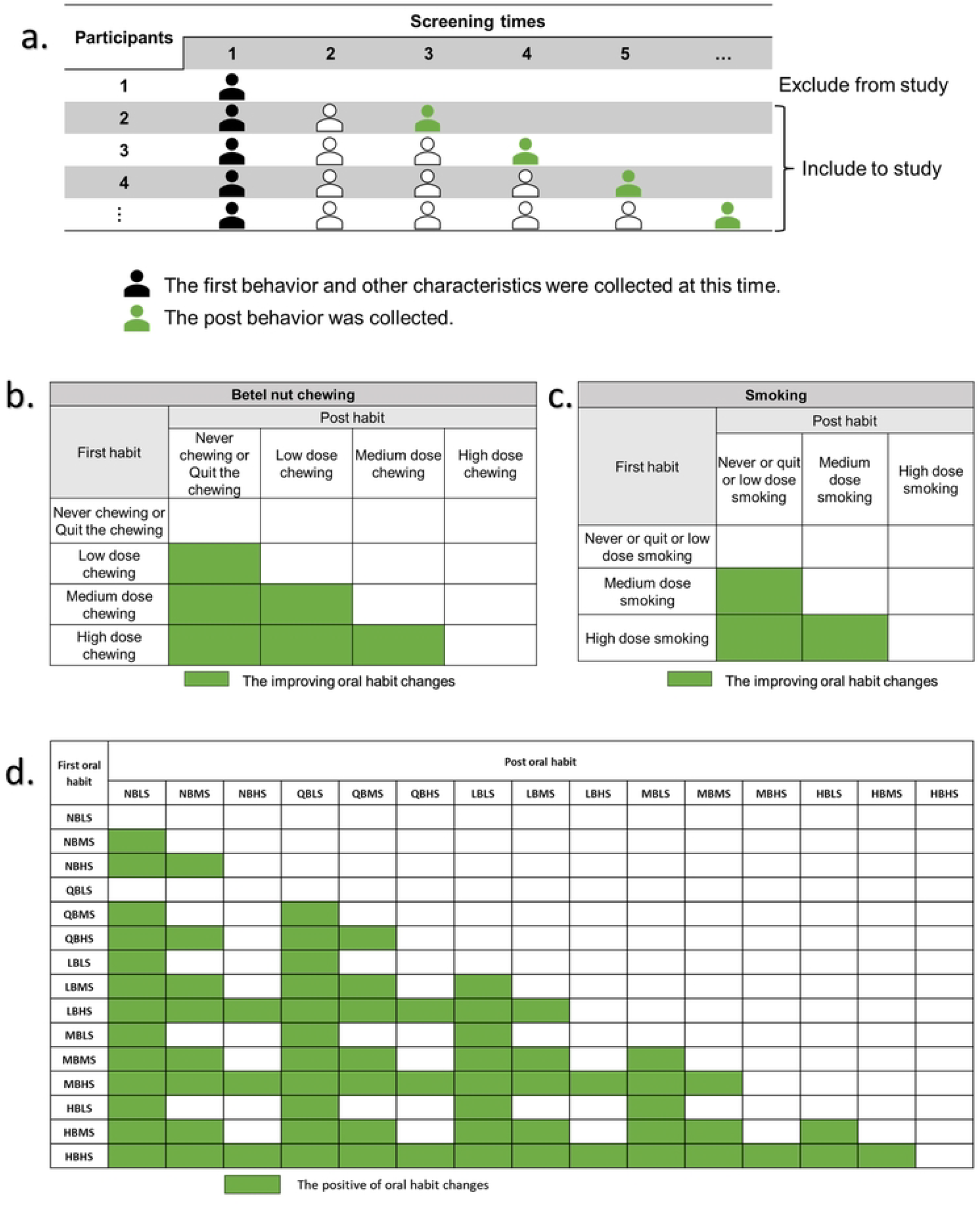
Participant Criteria and Definition of Behavioral Improvement in Oral Cancer Screening. (a.) participants who attended at least two times of screening will be included in this study. The oral habit change was compared between the first and last time of screening. (b.) the improving oral habit changes of betel quid chewing. (c.) the improving oral habit changes of smoking. (d.) the improvement of combined oral habit changes between cigarette smoking and betel quid chewing indicates at least 1 habit has to be improved with no other habit getting worse. Oral habits: NB-never chewing betel quid, QB-quit the chewing, LB-low degree chewing, MB-medium degree chewing, HB-high degree chewing, LS-never or quit or low degree smoking, MS-medium degree smoking, and HS-high degree smoking.

### 2.5 Statistical analysis

The oral habit changes (improved, no change, or deterioration) were summarized as numbers and percentages by the following characteristics: oral habits of the first screening, sex, age, education, screening times, and OPMD screening results. The investigation of the associated factors for improving habit changes was established with the logistic regression model. The outcome of the model was improvement or non-improvement. For missing data, this study categorized it as one level of the data, allowing for adjustments during the analysis. Note that individuals in LS and also in QB/LB at baseline were not included in the logistic regression analysis because there did not have a space to improve. All analyses were conducted with SAS version 9.4 (SAS Institute, Cary, NC, USA).

### 2.6 Ethical considerations

The Research Ethics Committee of the National Taiwan University Hospital approved this study under the annual Institutional Review Board (IRB) approval process and waived the requirement for informed consent in accordance with institutional review board regulations. This study was conducted from 2022 to 2024 under ethical approval numbers 202204036W, 202306056W, and 202403087W for each respective year. The study protocol was reviewed and approved by the Health Promotion Administration of the Taiwanese government, adhering to the ethical principles of the Declaration of Helsinki and its amendments.

## 3 Results

### 3.1 Oral habit changes distribution

There were 2,569,920 participants aged 30 years and older had at least two times of screening between 2010 and 2021. Among the participants who underwent repeated screening, 25.3% made a change to improve their oral habits. For those never chewed betel quids, participants with medium-degree and high-degree smoking changed to improving habits by 8.8% and 33.1%, respectively. Participants stopped chewing betel quids with medium-degree smoking, and high-degree smoking changed to improve habits by 24.1% and 46.1%, respectively. Low-degree chewing betel quids with no smoking, low-degree smoking, medium-degree smoking, and high-degree smoking all improved to 34.4%, 52.8%, and 69.9%, respectively. Medium-degree chewing betel quids with no smoking, low-degree smoking, medium-degree smoking, and high-degree smoking improved to 42.3%, 53.3%, and 74.2%, respectively. High-degree chewing betel quids with no smoking, low-degree smoking, medium-degree smoking, and high-degree smoking improved to 48.7%, 55.9%, and 72.7%, respectively. Males changed to improving habits at 26.2%, while females were at 20.8%. Participants aged 30-45 years, 46-60 years, and over 61 years changed to improving habits at 25.2%, 26.8%, and 23%, respectively. 25.5% of participants whose highest education was in elementary school changed to improving habits, 26.7% whose highest education was in middle school and higher changed to improving habits, and 22.6% whose highest education was higher than college. 35.9% of participants with a positive OPMD screening result changed to improving habits, while 24.5% of the negative group changed to improving habits. Only 2 times and more than 2 times participation in this oral screening program changed to improving habits (23.7% and 27.1%, respectively) (Table 1).

**Table 1.**
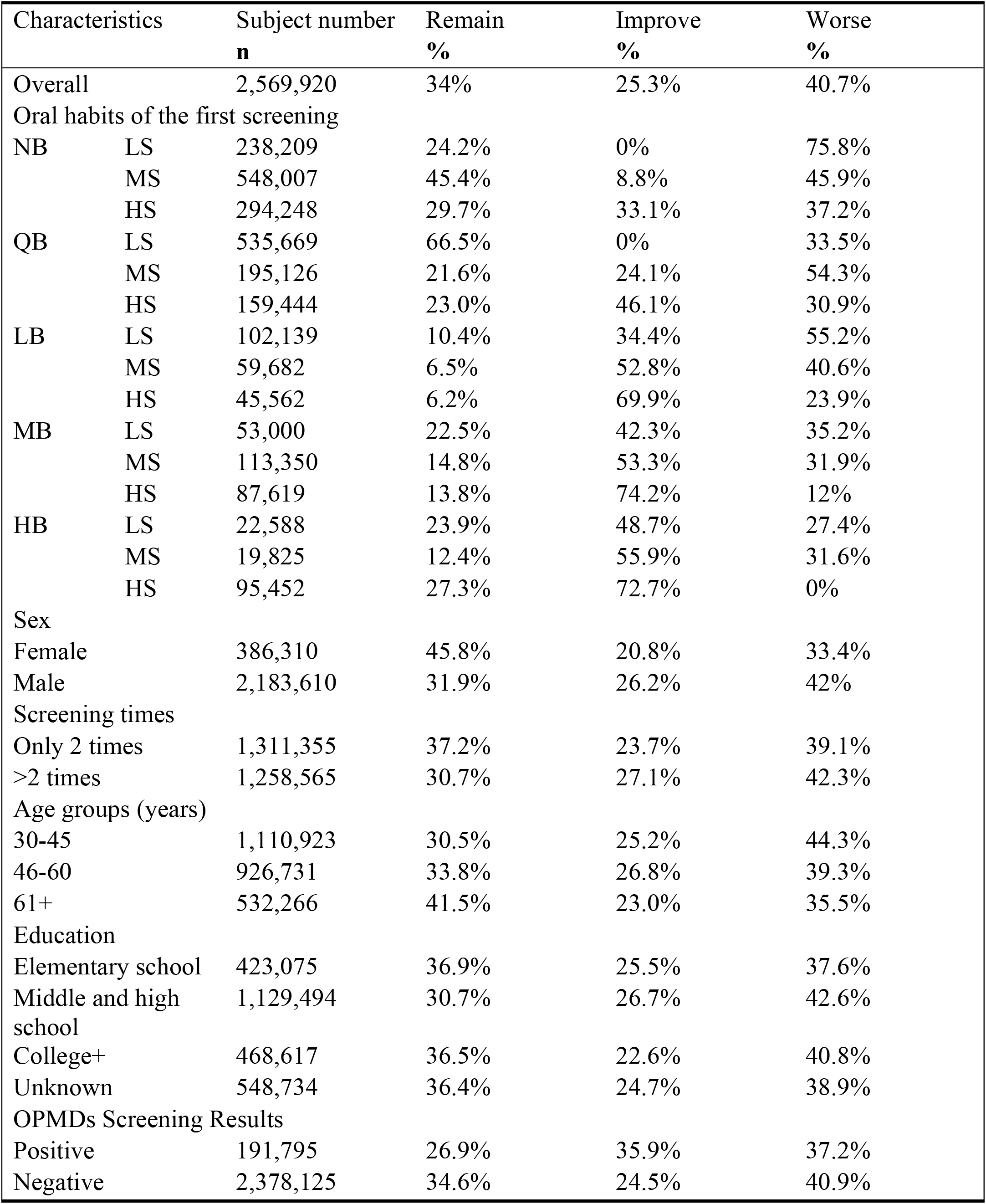
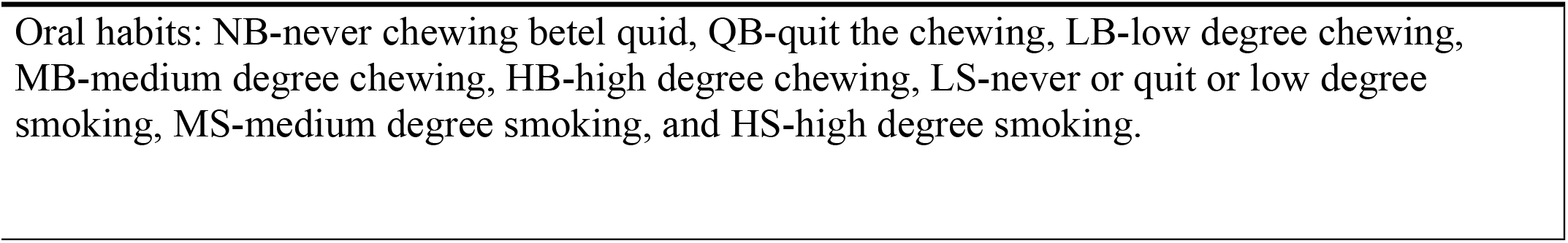
Characteristics of oral habit changes.

| Characteristics |  | Subject number<br>**n** | Remain<br>**%** | Improve<br>**%** | Worse<br>**%** |
| --- | --- | --- | --- | --- | --- |
| Overall |  | 2,569,920 | 34% | 25.3% | 40.7% |
| Oral habits of the first screening |  |  |  |  |  |
| NB | LS | 238,209 | 24.2% | 0% | 75.8% |
|  | MS | 548,007 | 45.4% | 8.8% | 45.9% |
|  | HS | 294,248 | 29.7% | 33.1% | 37.2% |
| QB | LS | 535,669 | 66.5% | 0% | 33.5% |
|  | MS | 195,126 | 21.6% | 24.1% | 54.3% |
|  | HS | 159,444 | 23.0% | 46.1% | 30.9% |
| LB | LS | 102,139 | 10.4% | 34.4% | 55.2% |
|  | MS | 59,682 | 6.5% | 52.8% | 40.6% |
|  | HS | 45,562 | 6.2% | 69.9% | 23.9% |
| MB | LS | 53,000 | 22.5% | 42.3% | 35.2% |
|  | MS | 113,350 | 14.8% | 53.3% | 31.9% |
|  | HS | 87,619 | 13.8% | 74.2% | 12% |
| HB | LS | 22,588 | 23.9% | 48.7% | 27.4% |
|  | MS | 19,825 | 12.4% | 55.9% | 31.6% |
|  | HS | 95,452 | 27.3% | 72.7% | 0% |
| Sex |  |  |  |  |  |
| Female |  | 386,310 | 45.8% | 20.8% | 33.4% |
| Male |  | 2,183,610 | 31.9% | 26.2% | 42% |
| Screening times |  |  |  |  |  |
| Only 2 times |  | 1,311,355 | 37.2% | 23.7% | 39.1% |
| >2 times |  | 1,258,565 | 30.7% | 27.1% | 42.3% |
| Age groups (years) |  |  |  |  |  |
| 30-45 |  | 1,110,923 | 30.5% | 25.2% | 44.3% |
| 46-60 |  | 926,731 | 33.8% | 26.8% | 39.3% |
| 61+ |  | 532,266 | 41.5% | 23.0% | 35.5% |
| Education |  |  |  |  |  |
| Elementary school |  | 423,075 | 36.9% | 25.5% | 37.6% |
| Middle and high school |  | 1,129,494 | 30.7% | 26.7% | 42.6% |
| College+ |  | 468,617 | 36.5% | 22.6% | 40.8% |
| Unknown |  | 548,734 | 36.4% | 24.7% | 38.9% |
| OPMDs Screening Results |  |  |  |  |  |
| Positive |  | 191,795 | 26.9% | 35.9% | 37.2% |
| Negative |  | 2,378,125 | 34.6% | 24.5% | 40.9% |

Oral habits: NB-never chewing betel quid, QB-quit the chewing, LB-low degree chewing, MB-medium degree chewing, HB-high degree chewing, LS-never or quit or low degree smoking, MS-medium degree smoking, and HS-high degree smoking.

Quit betel quid chewing with medium-degree betel quid chewing with high-degree smoking (MBHS) and high-degree betel quid chewing with high-degree smoking (HBHS) in males could improve habits by more than 70%, while low-degree betel quid chewing with high-degree smoking (LBHS), medium-degree betel quid chewing with high-degree smoking (MBHS), and high-degree betel quid chewing with high-degree smoking (HBHS) in females could improve habits by more than 70%. Positive OPMDs who engaged in medium-degree betel quid chewing with high-degree smoking (MBHS) and high-degree betel quid chewing with high-degree smoking (HBHS) saw an improvement in their habits of more than 70%. On the other hand, when the negative group engaged in low-degree betel quid chewing with high-degree smoking (LBHS), medium-degree betel quid chewing with high-degree smoking (MBHS), and high-degree betel quid chewing with high-degree smoking (HBHS), their habits improved by more than 70%. Improving habits more than 70% found in medium-degree betel quid chewing with high-degree smoking (MBHS) of aged 30-45 years; low-degree betel quid chewing with high-degree smoking (LBHS), medium-degree betel quid chewing with high-degree smoking (MBHS), and high-degree betel quid chewing with high-degree smoking (HBHS) of aged 46-60 years; low-degree betel quid chewing with smoking over medium-degree level (LBMS LBHS), medium-degree betel quid chewing with high-degree smoking (MBHS) and high-degree smoking with smoking over medium-degree level (HBMS HBHS) of aged over 61 years. Improving habit of more than 70% was found in low-degree betel quid chewing with high-degree smoking (LBHS), medium-degree betel quid chewing with high-degree smoking (MBHS) and high-degree betel quid chewing with high-degree smoking (HBHS) of participants whose highest education was elementary school; medium-degree betel quid chewing with high-degree smoking (MBHS) and high-degree betel quid chewing with high-degree smoking (HBHS) of participants whose highest education was middle and high school; low-degree betel quid chewing with high-degree smoking (LBHS), medium-degree betel quid chewing with high-degree smoking (MBHS) and high-degree betel quid chewing with high-degree smoking (HBHS) of participants whose highest education level was over college. Two-time screening participation resulted in an improvement of more than 70% in medium-degree betel quid chewing with high-degree smoking (MBHS); on the other hand, more than two-time screening participation led to an improvement of more than 70% in low-degree betel quid chewing with high-degree smoking (LBHS), medium-degree betel quid chewing with high-degree smoking (MBHS), and high-degree betel quid chewing with high-degree smoking (HBHS) (S1 Table).

Oral habits: NB-never chewing betel quid, QB-quit the chewing, LB-low degree chewing, MB-medium degree chewing, HB-high degree chewing, LS-never or quit or low degree smoking, MS-medium degree smoking, and HS-high degree smoking.

### 3.2 Factors related to better habit changes

The logistic regression model revealed that the majority of factors positively influenced the likelihood of enhancing habit improvements. To simplify the categorization of oral habits during the first round of screenings, we consolidate them into four distinct groups: only cigarette smoking, betel quid chewing combined with mild smoking, betel quid chewing combined with moderate smoking, and betel quid chewing combined with severe smoking. This approach helps to avoid an excessive number of levels. The study revealed that the favorable results of OPMDS had a significant impact on improving habits, but only in people who had a smoking habit exclusively (adjusted odds ratio (aOR) 1.18, 95% confidence interval (CI) 1.16–1.20). However, the presence of OPMD did not lead to improvement in habits for individuals who engaged in both smoking and chewing betel quids (aOR 0.84 (95% CI 0.82-0.87) for individuals who chewed betel quid and engaged in mild smoking, aOR 0.79 (95% CI 0.77-0.81) for those who chewed Betel quid and engaged in moderate smoking, aOR 0.88 (95% CI 0.86-0.9) for those who chewed betel quid and engaged in severe smoking). Males were less likely to improve their oral habits compared to females (aOR = 0.86, 95% CI: 0.85-0.87). Compared to individuals aged 30-45 years, it demonstrated that as age increased, there was a significant increase in improving oral habit. The adjusted odds ratio (95% confidence interval) was 1.31 (1.3–1.32) for individuals aged 46–60 years and 1.71 (1.69–1.73) for those over 61 years. Those with the highest education level in middle and high school showed a decreased likelihood of habit improvement when comparing their education to the elementary school level (aOR 0.94, 95% CI 0.93-0.95), while individuals with the highest level of education beyond college have a greater chance of improving their habits (aOR 1.09, 95% CI 1.08–1.11). Participating attending more than two rounds of screening, compared to two rounds of screening, significantly increased the opportunity for habit improvement (aOR 1.046, 95% CI 1.038-1.053) (Table 2).

**Table 2.** The associated factors of improving habit changes.

| Characteristics | OR | 95% CI |  | aOR | 95% CI |  |
| --- | --- | --- | --- | --- | --- | --- |
| Oral habits of the first screening \| OPMD results |  |  |  |  |  |  |
| Only cigarette smoking |  |  |  |  |  |  |
| Positive OPMD | 1.18 | 1.16 | 1.2 | 1.18 | 1.16 | 1.2 |
| Negative OPMD | ref |  |  | ref |  |  |
| Betel quid chewing with mild smoking |  |  |  |  |  |  |
| Positive OPMD | 0.82 | 0.79 | 0.85 | 0.84 | 0.82 | 0.87 |
| Negative OPMD | ref |  |  | ref |  |  |
| Betel quid chewing with moderate smoking |  |  |  |  |  |  |
| Positive OPMD | 0.78 | 0.76 | 0.8 | 0.79 | 0.77 | 0.81 |
| Negative OPMD | ref |  |  | ref |  |  |
| Betel quid chewing with severe smoking |  |  |  |  |  |  |
| Positive OPMD | 0.88 | 0.86 | 0.9 | 0.88 | 0.86 | 0.9 |
| Negative OPMD | ref |  |  | ref |  |  |
| Sex |  |  |  |  |  |  |
| Male | 1.19 | 1.17 | 1.2 | 0.86 | 0.85 | 0.87 |
| Female | ref |  |  | ref |  |  |
| Age groups (years) |  |  |  |  |  |  |
| 30-45 | ref |  |  | ref |  |  |
| 46-60 | 1.17 | 1.16 | 1.18 | 1.31 | 1.3 | 1.32 |
| 61+ | 1.3 | 1.29 | 1.31 | 1.71 | 1.69 | 1.73 |
| Education |  |  |  |  |  |  |
| Elementary school | ref |  |  | ref |  |  |
| Middle and high school | 0.83 | 0.82 | 0.84 | 0.94 | 0.93 | 0.95 |
| College+ | 0.684 | 0.677 | 0.692 | 1.09 | 1.08 | 1.11 |
| Screening time |  |  |  |  |  |  |
| Screening 2 times | ref |  |  | ref |  |  |
| Screening more than 2 times | 1.086 | 1.08 | 1.093 | 1.046 | 1.038 | 1.053 |

## 4 Discussion

In this study, we demonstrate a positive correlation between smoking intensity and habit enhancement, whereas betel quid chewing intensity did not influence habit improvement based on a large-scaled oral cancer screening program targeting subjects with habits of smoking/betel quid chewing. High-degree smoking influences improve oral risk habits, especially in high-degree smokers with medium- and high-degree betel quid chewing. This finding was similar to a Malaysian study’s, which indicated that those who consumed both betel quids and tobacco face more challenges when attempting to quit betel quid consumption [8]. However, US and Korean populations showed that people with high-degree smoking decreased the opportunity for improving oral risk habits (smoking cessation), with an OR (95% CI) of 0.42 (0.35, 0.5) and 0.67 (0.47, 0.95), respectively [12, 13]. This study considered reducing the frequency of smoking or chewing betel quids as a form of enhanced habit, while other studies concentrated on cessation, potentially explaining the disparity in outcomes.

Those diagnosed with OPMDs can increase health awareness and promote habit modification among cigarette smokers. The study in China and Korea, which found that participant illness influenced smoking improvement [13, 22], supports this result. According to a study in China, illness was the 47.3% most common reason for smoking cessation [22]. In Korea, a study found that illness affected smoking cessation with an OR (95% CI) of 1.4 (1.07, 1.85) for hypertension and 1.68 (1.03, 2.75) for cardiovascular disease [13]. Some suspected abnormal findings on CT screening affected smoking’s improvement [23, 24]. However, a positive diagnosis for OPMDs does not have an impact on habit improvement in individuals who both smoke cigarettes and chew betel quids. Evidently, the habit of chewing betel quids is more resistant to improvement than smoking, despite the presence of disease risk indicators.

Male individuals demonstrated a greater capacity for enhancing their habits in comparison to their female counterparts. However, consideration of other variables revealed that males showed less improvement in their habits than females. Previous research in the US and China, with adjusted odds ratios (aOR) of 0.96 (95% CI: 0.84–1.11) for the US [12] and 0.81 (95% CI: 0.43– 1. 54) for China [14], also found that men were less likely to quit smoking. Conversely, the Malaysian study found a hazard rate ratio (HRR) of 0.82 for females who stopped chewing betel quids, with a 95% confidence interval spanning from 0.5 to 1.3 [8]. Nevertheless, the outcomes of all three trials did not demonstrate statistical significance. In China, further research showed a similar distribution, with 16.6% of males and 16.3% of females intending to quit smoking [22]. Evidence indicates that both males and females can contribute to habit enhancement, with differing effects according on geographical area.

The opportunity for improving the oral risk habits of both models in this study is higher as age increases. However, researchers found that elderly Malaysians were less likely to stop their betel quid chewing habit (HRR 0.2, 95% CI 0.1-0.6) [8]. Additionally, some studies of smoking cessation in China and Korea showed a non-significant effect of age, with an OR (95% CI) of 0.94 (0.4, 2.23) and 2.35 (0.48, 11.45), respectively [11, 14]. The disparities in oral health outcomes among areas can be ascribed to variations in health awareness and compliance with traditional practices.

The participants’ highest level of education, particularly a college degree or higher, can positively influence habit improvement. The findings of this study align with prior research conducted in China and Korea. In China, a study found that education has a non-significant effect on improving cigarette smoking, with an OR (95% CI) of 1.21 (0.84, 1.74) for medium education and 1.29 (0.83, 1.98) for high education, compared to low education. However, this survey study in China was limited to urban areas, but most Chinese smokers live in rural areas [14]. The study in Korea showed that high education increases the opportunity for improving cigarette smoking, with an OR (95% CI) of 3.19 (1.02, 9.98) [11]. Highly educated persons sometimes prioritize their personal opinions over societal conventions. Once individuals comprehensively grasp the risks linked to their behaviors, they are more inclined to implement modifications.

Conducting screenings more than twice could improve participants’ habits. Repeated participation indicates a deliberate effort to change these oral risky habits and maintain a healthy lifestyle. Other studies in China and Korea also found the same pattern: participants who intended to quit smoking were more successful. The study in Korea used the clinic visit period to demonstrate the intention to quit smoking and found the impact to be an OR (95% CI) of 7.16 (5.57, 9.2) [13]. Among Korean participants who consulted about smoking cessation, counseling increased the opportunity for success, with an OR (95% CI) of 1.87 (1.21, 2.89) [11]. Among Chinese participants, those who tried to quit smoking had more improvement of smoking cessation, with an OR (95% CI) of 2.29 (1.81, 2.89) [14].

The investigation reveals that a quarter of the participants in an oral cancer screening program were able to effectively enhance their practices. The screening program offers both early diagnosis of abnormal lesions or diseases and knowledge about oral risk factors that can lead to oral cancer. Research suggests that individuals who heavily consume betel quids and smoke, especially those with a smoking addiction and those diagnosed with oral potentially malignant disorders (OPMD), can effectively alter their habit. OPMDs, or Oral Potentially Malignant Disorders, encompass a range of abnormalities that have the potential to progress into oral cancer. Consequently, upon discovery, individuals enhance their awareness and refine their actions. Based on the principles of nature, the human body undergoes degradation over time, which could explain why the older individuals involved in the study possess a greater understanding of health risks compared to the younger ones. Consequently, the elderly exhibit positive cooperation in enhancing their habits. People consider a college education or higher to be an advanced level of education. These individuals prefer to stick to their own concepts rather than social values. Once they comprehend the hazards of their habits, they decide to adjust them. Consistent participation in the screening program demonstrates a genuine and resolute commitment to health well-being, leading to the successful cultivation of healthier habits. Furthermore, oral professionals such as doctors, nurses, and technicians, who have undergone specialized training, administer the oral cancer screening program in Taiwan. This ensures the program’s credibility. These specialists are probably accountable for the program’s efficacy in fostering beneficial habits [25–30].

## 4.1 Limitations

Nevertheless, we used participant cognition to gather data on their highest level of education and information about cigarette or betel quid consumption. As a result, the occurrence of recall bias is a very plausible scenario. The current study had one limitation. As we had a particular set of people who participated in the screening program at least twice as the subject of the current study, the results could not be generalized to people who participated in the screening program only once.

## 5 Conclusion

This study highlights that 25% of participants improved their oral habits, reflecting a positive outcome of the oral cancer screening program. The most significant improvements were observed among elderly individuals, those with higher education levels, and participants who consistently engaged with the program. These findings underline the potential of such initiatives to foster healthier habits, particularly among groups more likely to be influenced by health education and regular monitoring. However, being diagnosed with oral potentially malignant disorders (OPMDs) has not proven to be a strong motivator for habital change, except among individuals who smoked but did not engage in other risky habits, such as betel quid chewing. This indicates a gap in awareness among high-risk groups engaging in multiple risky habits, who seem less likely to recognize the dangers of their habits or the severity of OPMDs.

To address this issue, future efforts should focus on enhancing public knowledge and understanding through collaboration between health organizations and the screening program. By promoting greater awareness of the risks associated with smoking, betel quid chewing, and OPMDs, such partnerships could help individuals recognize the importance of changing their habits. Increased understanding could empower high-risk groups to make more informed decisions about their health. Enhancing the effectiveness of the oral cancer screening program could further reduce the incidence of oral cancer over time and significantly lower the healthcare costs associated with its treatment. These combined efforts would not only improve individual outcomes but also strengthen the overall impact of the screening initiative on public health.

## Supporting information

S1 Table. Table Characteristics of oral habit change in model (improve and not improve)

## Acknowledgments

The authors provide their warm thanks and appreciation to all study participants.

## Author contributions

All authors discussed the results and contributed to the final manuscript. PM: Writing – original draft, Formal analysis, Software. CWS: Data curation, Methodology. HFY and THH: Project administration, Investigation. YYC, LJL, and CCW: Project administration, Funding acquisition. SLSC: Methodology, Supervision. AMFY: Conceptualization, Methodology, Supervision, Validation, Writing – review & editing.

## Data availability statement

The raw data supporting the conclusions of this article will be made available by the authors without undue reservation.

